# Immune response to SARS-CoV-2 in the nasal mucosa in children and adults

**DOI:** 10.1101/2021.01.26.21250269

**Authors:** Clarissa M Koch, Andrew D Prigge, Kishore R Anekalla, Avani Shukla, Hanh Chi Do-Umehara, Leah Setar, Jairo Chavez, Hiam Abdala-Valencia, Yuliya Politanska, Nikolay S Markov, Grant R Hahn, Taylor Heald-Sargent, L Nelson Sanchez-Pinto, William J Muller, Alexander V Misharin, Karen M Ridge, Bria M Coates

## Abstract

**Rationale:** Despite similar viral load and infectivity rates between children and adults infected with SARS-CoV-2, children rarely develop severe illness. Differences in the host response to the virus at the primary infection site are among the proposed mechanisms.

**Objectives:** To investigate the host response to SARS-CoV-2, respiratory syncytial virus (RSV), and influenza virus (IV) in the nasal mucosa in children and adults.

**Methods:** Clinical outcomes and gene expression in the nasal mucosa were analyzed in 36 children hospitalized with SARS-CoV-2 infection, 24 children with RSV infection, 9 children with IV infection, 16 adults with mild to moderate SARS-CoV-2 infection, and 7 healthy pediatric and 13 healthy adult controls.

**Results:** In both children and adults, infection with SARS-CoV-2 leads to an interferon response in the nasal mucosa. The magnitude of the interferon response correlated with the abundance of viral reads and was comparable between symptomatic children and adults infected with SARS-CoV-2 and symptomatic children infected with RSV and IV. Cell type deconvolution identified an increased abundance of immune cells in the samples from children and adults with a viral infection. Expression of *ACE2* and *TMPRSS2* – key entry factors for SARS-CoV-2 – did not correlate with age or presence or absence of viral infection.

**Conclusions:** Our findings support the hypothesis that differences in the immune response to SARS-CoV-2 determine disease severity, independent of viral load and interferon response at the primary infection primary site.

## Introduction

Severe acute respiratory syndrome coronavirus 2 (SARS-CoV-2) is responsible for the current pandemic known as coronavirus disease 2019 (COVID-19). A notable feature of SARS-CoV-2 infection is the dramatic heterogeneity in clinical phenotypes. While most infected individuals remain asymptomatic or develop a very mild disease, some develop severe SARS-CoV-2 pneumonia and acute respiratory distress syndrome. These severe infections account for most of the public health and societal impacts of the COVID-19 pandemic and disproportionately affect the elderly while mostly sparing children (1–3). This is different from other common viral respiratory infections, including respiratory syncytial virus (RSV) and influenza virus (IV), which tend to be most severe in young children and elderly adults. Of the approximately 225,000 deaths from COVID-19 reported in the first eight months of the pandemic in the United States, only 189 occurred in children less than 18 years of age (4), despite at least 277,285 documented infections in children over the same period (5).

Identifying the mechanisms driving morbidity and mortality differences between children and adults infected with SARS-CoV-2 may provide essential insights for treating patients with COVID-19. Infection begins in the nasal mucosa (6, 7), where the SARS-CoV-2 spike protein binds angiotensin-converting enzyme 2 (ACE2) on the surface of epithelial cells. In concert with host proteases, principally transmembrane serine protease 2 (TMPRSS2), the virus enters the cell to begin replication (8–10). Multiple studies demonstrated that *ACE2* expression in the airways increases with age (11, 12), leading to lower infectivity and lower viral load in children. However, subsequent studies have demonstrated that SARS-CoV-2 viral load is similar in children and adults, suggesting the relative protection from COVID-19 in children is not due to age-related differences in infectivity (13–16). As there are important differences between the pediatric and adult immune systems, the host response to SARS-CoV-2 could explain the observed differences in n children and adults’ clinical phenotypes. The primary host response to viral infection is antiviral interferon signaling to limit viral replication, recruit immune cells, and clear infected cells (17). However, some viruses have developed strategies to evade this response, and SARS-CoV-2 is known to inhibit host interferon responses *in vitro (18, 19)*. This has led investigators to speculate that differences in antiviral interferon signaling at the primary infection site might be responsible for differences in the frequency of severe COVID-19 between children and adults. To date, no studies have compared the host response to the virus at the primary site of SARS-CoV-2 infection in children and adults.

This study aimed to assess how the host response to SARS-CoV-2 at the primary site of infection compares between children and adults, and how it may differ from the host response to other respiratory viruses, namely R.S.V. and IV, that are known to cause severe disease in children. Curettage and transcriptional profiling of the nasal mucosa can provide valuable insights into host-pathogen interactions at the initial infection site that may drive disease outcomes (20–23). Therefore, we performed R.N.A. sequencing of nasal mucosa samples from children and adults with SARS-CoV-2, children with R.S.V. or IV, and healthy children and adults. Local antiviral interferon signaling did not differ between children and adults infected with SARS-CoV-2 and was similar to children with severe R.S.V. infection. The magnitude of the interferon response only correlated with viral load and not the severity of illness. Our findings suggest that the local interferon response to SARS-CoV-2 in the upper respiratory tract does not differ between children and adults, arguing that factors other than the local antiviral response drive differences in the clinical phenotypes associated with SARS-CoV-2 infection.

## Methods

### Study population

Our study included the following six groups: healthy children, children infected with SARS-CoV-2, children infected with RSV, children infected with IV, healthy adults, and adults infected with SARS-CoV-2. Approval for this study was obtained from the Institutional Review Board at Ann & Robert H. Lurie Children’s Hospital of Chicago (Chicago, IL, U.S.A.). Informed consent was obtained from adult participants and parents/guardians of pediatric participants. The medical records of all pediatric patients admitted between December 2019, and November 2020 were screened for inclusion criteria and the absence of exclusion criteria (see below). We sequenced samples from healthy children recruited for a previous study as negative controls. These samples were obtained from healthy children attending a well-child check at Ann & Robert H. Lurie Children’s Hospital of Chicago Outpatient Center in May 2018. If the patient was eligible for the study, a study team member approached for consent. For SARS-CoV-2-infected children, both the child and accompanying adult were invited to participate.

### Inclusion criteria

Children were eligible for inclusion in the SARS-CoV-2-infected group if they had a positive polymerase chain reaction (PCR) test for SARS-CoV-2 at admission to the hospital or our drive through testing center (**Supplemental Figure 1A**). Symptoms were collected based on reports by participants or their guardians.

Adults accompanying a SARS-CoV-2-infected child or who were themselves admitted and cared for in our pediatric hospital with a positive PCR test for SARS-CoV-2 were eligible for inclusion (**Supplemental Figure 1B**). Polymerase chain reaction testing was not available for the adults accompanying a child prior to enrollment. RNA sequencing identified SARS-CoV-2 RNA in adult participants categorized with SARS-CoV-2. Adults without detectable SARS-CoV-2 via RNA sequencing were classified as healthy controls. Self-reported symptoms were collected from adults at the time of enrollment and via follow-up weekly phone calls for 3 weeks. Adults were asked about 11 of the most common symptoms reported with SARS-CoV-2 infection.

Participants were eligible for inclusion in the positive viral control group if they had a PCR confirming infection with RSV or IV, were admitted to the pediatric intensive care unit (PICU), and had symptoms of pulmonary parenchymal disease, including any of the following: 1) infiltrates, areas of atelectasis, or hyperinflation on chest radiograph, 2) rales, wheezing, retractions, tachypnea, or any form of respiratory distress on clinical exam, or 3) an oxygen requirement to maintain peripheral oxygen saturation levels above 95%.

Healthy pediatric controls were eligible for inclusion if they were less than 18 years of age and had no signs or symptoms of viral infection at the time of their scheduled well-child visit with their Primary Care Provider (**Supplemental Figure 1C**). All healthy pediatric controls were clinically stable and without apparent viral or bacterial infection when the nasal epithelial scrapings were obtained. Additionally, metagenomic analysis of RNA-seq data using the OneCodex platform did not detect any viral reads for 29 common human respiratory viruses. Healthy controls were followed for 2 weeks via electronic medical record to ensure no infections occurred within that time frame.

### Exclusion criteria

Subjects were not eligible for inclusion in the pediatric healthy control group if they had any significant medical co-morbidities, including pre-existing lung disorders, pre-existing cardiac disease, immunodeficiency, malignancy, and neurologic disorders, increasing the risk of aspiration or respiratory failure. Children with a clinical concern for multisystem inflammatory syndrome in children (MIS-C) were not eligible for inclusion in the SARS-CoV-2 cohort.

### Patient Enrollment

We enrolled 163 participants in the study (**Supplemental Figure 1**). Prior to sample collection, 10 participants withdrew consent. Eight participants were discharged before sample collection. After the isolation of RNA, 26 samples were excluded due to low RNA quality. Following RNA sequencing, an additional 14 samples were excluded: 1 for low-quality RNA-seq library, 3 for sex mismatch between biological sex indicated in the research records and expression of *XIST* or *RPS4Y1* genes, 3 for negative PCR testing results, 1 for multisystem inflammatory syndrome in children (MIS-C) diagnosis, 1 for unavailable patient research records, and 5 healthy controls for detection of viral reads for common respiratory viruses. A cohort of 105 participants: 7 healthy children, 36 children infected with SARS-CoV-2, 24 children infected with RSV, 9 children infected with IV, 13 healthy adults, and 16 adults infected with SARS-CoV-2 were used for the analysis.

### Clinical Data

Clinical data were collected and managed using REDCap electronic data capture tools hosted at the Northwestern University Clinical and Translational Sciences Institute (24). Hospital length of stay was determined from admission and discharge dates. Readmissions within 24 hours of discharge were considered a continuation of hospitalization. If a patient had multiple PICU stays during a single encounter, the PICU length of stay was cumulative. Advanced respiratory support was defined by the use of high flow nasal cannula (HFNC), non-invasive positive pressure ventilation (NIPPV), or invasive mechanical ventilation (IMV). Respiratory support and oxygen duration were determined by documentation on a calendar day of non-home advanced respiratory support or oxygen to exclude chronic baseline support. Pediatric acute respiratory distress syndrome was defined using the Pediatric Acute Lung Injury Consensus Conference criteria (25). Detection of a co-pathogen was defined as a positive viral PCR or any positive bacterial or fungal cultures.

### Nasal curettage and RNA isolation

Due to infection control procedures during the COVID-19 pandemic, nasal mucosa curettage was performed during routine care by health care providers caring for the patient. Providers received training just before sample collection by trained study personnel. Nasal epithelial cells were obtained by mucosal scrape biopsy of the inferior turbinate using a sterile plastic curette (Rhino-Pro curette, Arlington Scientific) as previously described (26) to obtain a predominantly epithelial cell population. A total of 3 single-pass scrapings per nares were performed per subject. The curette tips were cut and placed into RNase-free collection tubes containing 350 μL of RLT plus buffer (Qiagen, Germantown, MD, U.S.A.) supplemented with 2-mercaptoethanol, vortexed vigorously, and stored at −80°C. RNA was isolated using Qiagen RNeasy extraction kits (Qiagen).

### RNA sequencing and analysis

RNA quality and quantity were assessed using TapeStation 4200 High Sensitivity RNA tapes (Agilent), and RNA-seq libraries were prepared from 1 ng of total RNA using SMARTer Stranded Total RNA-seq Kit v2 (Takara Bio). After QC using TapeStation 4200 High Sensitivity DNA tapes (Agilent), dual indexed libraries were pooled and sequenced on a NextSeq 500 instrument (Illumina), 75 cycles, single-end, to an average sequencing depth of 5.88M reads.

FASTQ files were generated using bcl2fastq (Illumina). Viral RNA was detected using a custom hybrid genome prepared by joining FASTA, GFF, and GTF files for GRCh37.87, SARS-CoV-2 (NC_045512.2), Influenza A/California/07/2009 (GCF_001343785.1), and RSV/S2 ts1C (GCF_000856445.1) genomes. An additional negative-strand transcript spanning the entirety of the SARS-CoV-2 genome was then added to the GTF and GFF files to enable detection of SARS-CoV-2 replication. To facilitate reproducible analysis, samples were processed using the publicly available nf-core/RNA-seq pipeline version 1.4.2 implemented in Nextflow 19.10.0 using Singularity 3.2.1-1 with the minimal command `nextflow run nf-core/rnaseq -r 1.4.2 –singleEnd -profile singularity –reverseStranded --three_prime_clip_r2 3` (27–29). Briefly, lane-level reads were trimmed using trimGalore! 0.6.4 and aligned to the hybrid genome described above using STAR 2.6.1d (30). Gene-level assignment was then performed using featureCounts 1.6.4 (31). First, putative sample swaps were identified by comparing known patient sex with sex determined by *XIST* and *RPS4Y1* expression levels. No compromised samples were not included in the analysis.

### Weighted gene co-expression network analysis (WGCNA)

WGCNA was performed manually using WGCNA version 1.69 with default settings unless otherwise noted (32). Highly variable genes among all participant samples (n=105) and pediatric participant samples only (n=76) were included in analyses. To best capture patterns of co-regulation, a signed network was used. Using the `pickSoftThreshold` function, we empirically determined a soft threshold of 3 to best fit the network structure. A minimum module size of 30 genes was chosen. Module eigengenes were then related to patient and sample metadata using biweight midcorrelation. Module GO enrichment was then determined using GOrilla (33).

### Deconvolution of bulk RNA-seq signatures

Deconvolution of bulk RNA-seq signatures was performed using AutoGeneS v1.0.3 (34). For details of the functions used and their parameters, see the code available at https://github.com/NUPulmonary/2021_Koch. Briefly, we used a single-cell RNA-seq dataset from Ordovas-Montanes et al. (35), which contains nasal epithelial and immune cells, to train the AutoGeneS model. AutoGeneS selects cell type-specific genes by both minimizing the correlation between and maximizing the distance between the cell type-specific average gene expression profiles. The model was then applied to bulk RNA-seq data to estimate specific cell types’ proportions using regression.

### Statistical Analysis

Categorical data were summarized by percentages and compared using Fisher’s exact test. Continuous, non-parametric data were summarized using medians with interquartile ranges and compared using Mann-Whitney U or Kruskal-Wallis testing. The Benjamini-Hochberg procedure was used for false discovery rate correction when performing multiple comparisons. Data were considered statistically significant at P-adjusted<0.05. Statistical analyses were performed using Graphpad Prism version 9.0.0 and R version 4.0.3. Data were visualized using ggplot2 version 3.3.1 and Graphpad Prism.

### Data and code availability

Processed RNA-seq counts are available as Online Supplemental Table 1. Raw data is in the process of being deposited to dbGaP/SRA. Code is available via https://github.com/NUPulmonary/2021_Koch.

## Results

### Description of the cohort

The demographics of the pediatric and adult cohorts are summarized in **Table 1** and **Supplemental Table 1**. When considering only children, patients with RSV were younger, and a larger proportion was previously healthy than those with SARS-CoV-2 (**Table 1**). The ages and proportion of individuals with chronic medical conditions in the uninfected adult group were no different from the SARS-CoV-2 infected adult group (**Supplemental Table 1**).

**Table 1:** Description of Pediatric Cohort. Adjusted p-value <0.05 were considered statistically significant, calculated by Kruskal-Wallis test with Benjamini-Hochberg FDR correction.

|  | **SARS-CoV-2**<br>**Children** | **RSV**<br>**Children** | **IV**<br>**Children** | **P-Adj** |
| --- | --- | --- | --- | --- |
| **n** | 36 | 24 | 9 |  |
| **Median Age (yrs) [IQR]** | 1.9 [0.4-15.0] | 0.33 [0.16-0.44] | 1.7 [1.4-7.0] | 0.003 |
| **Sex** |  |  |  |  |
| Male (%) | 18 (50.0) | 10 (41.7) | 5 (55.6) | 0.92 |
| Female (%) | 18 (50.0) | 14 (58.3) | 4 (44.4) |  |
| **Race** |  |  |  |  |
| White (%) | 8 (22.2) | 13 (54.2) | 1 (11.1) | 0.03 |
| African American (%) | 4 (11.1) | 1 (4.2) | 2 (22.2) |  |
| Asian (%) | 0 (0.0) | 2 (8.3) | 1 (11.1) |  |
| Other (%) | 23 (63.9) | 6 (25.0) | 4 (44.4) |  |
| Unspecified (%) | 1 (2.8) | 2 (8.3) | 1 (11.1) |  |
| **Ethnicity** |  |  |  |  |
| Non-Hispanic (%) | 11 (30.6) | 15 (62.5) | 4 (44.4) | 0.14 |
| Hispanic (%) | 24 (66.7) | 7 (29.2) | 4 (44.4) |  |
| Unspecified (%) | 1 (2.8) | 2 (8.3) | 1 (11.1) |  |
| **Medical History** |  |  |  |  |
| Healthy (%) | 12 (33.3) | 19 (79.2) | 3 (33.3) | 0.006 |
| Respiratory (%) | 9 (25.0) | 2 (8.3) | 5 (55.6) | 0.06 |
| Cardiovascular (%) | 5 (13.9) | 1 (4.2) | 1 (11.1) | 0.82 |
| Neurologic (%) | 6 (16.7) | 1 (4.2) | 2 (22.2) | 0.47 |
| Hematologic (%) | 0 (0.0) | 1 (4.2) | 0 (0) | 0.76 |
| Oncologic (%) | 2 (5.6) | 0 (0) | 0 (0) | 0.88 |
| Chronic Kidney Disease (%) | 1 (2.8) | 1 (4.2) | 0 (0) | 1 |
| Gastrointestinal (%) | 8 (22.2) | 2 (8.3) | 2 (22.2) | 0.62 |
| Endocrine (%) | 3 (8.3) | 2 (8.3) | 1 (11.1) | 1 |
| Metabolic/Genetic (%) | 8 (22.2) | 1 (4.2) | 2 (22.2) | 0.38 |
| **Presentation\*** |  |  |  |  |
| Respiratory (%) | 20 (57.1) | 24 (100) | 9 (100) | - |
| Gastrointestinal (%) | 3 (8.6) | 0 (0) | 0 (0) | - |
| Fever Only (%) | 5 (14.3) | 0 (0) | 0 (0) | - |
| Non-Respiratory Viral Syndrome (%) | 2 (5.7) | 0 (0) | 0 (0) | - |
| Asymptomatic (%) | 5 (14.3) | 0 (0) | 0 (0) | - |
| Median Day of Illness at Collection [IQR] | 4.0 [2.0-6.8] | 4.0 [3.0-4.3] | 5.0 [4.0-5.0] | 0.88 |
| **Severity** |  |  |  |  |
| ICU Admission (%) | 8 (22.2) | 24 (100) | 9 (100) | - |
| Median ICU LoS (d) [IQR] | 3.0 [2.0-6.0] | 7.0 [4.0-10.0] | 6.0 [3.0-9.0] | 0.68 |
| Median Hospital LoS (d) [IQR] | 3.0 [2.0-6.0] | 7.0 [5.0-12.0] | 6.0 [3.0-9.0] | 0.006 |
| ARDS (%) | 1 (2.8) | 3 (12.5) | 1 (11.1) | 0.62 |
| Vasoactive Medications | 1 (2.8) | 2 (8.3) | 0 (0) | 0.89 |
| **Respiratory Support** |  |  |  |  |
| *Collection* |  |  |  |  |
| None (%) | 29 (80.6) | 1 (4.2) | 0 (0) | 3.7E-13 |
| Low Flow Interface (%) | 4 (11.1) | 0 (0) | 0 (0) |  |
| HFNC (%) | 1 (2.8) | 13 (54.2) | 3 (33.3) |  |
| NIPPV (%) | 2 (5.6) | 1 (4.2) | 4 (44.4) |  |
| IMV (%) | 0 (0.0) | 9 (37.5) | 2 (22.2) |  |
| Oxygen (%) | 7 (19.4) | 23 (95.8) | 9 (100) | 1 |
| *Peak* |  |  |  |  |
| None (%) | 26 (72.2) | 0 (0) | 0 (0) | 1.4E-11 |
| Low Flow Interface (%) | 3 (8.3) | 0 (0) | 0 (0) |  |
| HFNC (%) | 3 (8.3) | 12 (50.0) | 3 (33.3) |  |
| NIPPV (%) | 4 (11.1) | 2 (8.3) | 4 (44.4) |  |
| IMV (%) | 0 (0.0) | 10 (41.7) | 2 (22.2) |  |
| Oxygen (%) | 9 (25.0) | 24 (100) | 9 (100) | 1 |
| Median Respiratory Support Days<sup>†</sup> [IQR] | 5.0 [2.0-6.0] | 6.5 [4.0-11.0] | 5.0 [3.0-8.0] | 0.62 |
| Median Supplemental Oxygen Days [IQR] | 3.0 [2.0-6.0] | 7.0 [4.8-10.0] | 5.0 [3.0-9.0] | 0.26 |
| **Co-Pathogen Detection** |  |  |  |  |
| Virus | 0 (0.0) | 3 (12.5) | 1 (11.1) | 0.17 |
| Bacteria | 0 (0.0) | 6 (25.0) | 1 (11.1) | 0.02 |

*Abbreviations:* P-adj=adjusted p value, IQR=interquartile range, ICU=intensive care unit, LoS=Length of stay, ARDS=acute respiratory distress syndrome, HFNC=High flow nasal canula, NIPPV=non-invasive positive pressure ventilation, IMV=invasive mechanical ventilation
\*n=35 for SARS-CoV-2 presentation due to incomplete medical records
<sup>†</sup>Cumulative days requiring HFNC, NIPPV, or IMV

**Figure 1A** summarizes the clinical course of the hospitalized pediatric participants infected with IV, RSV, or SARS-CoV-2. There was no difference in the time from symptom onset to nasal sample collection between the groups (**Figure 1B**). Consistent with their inclusion as positive controls for moderate and severe viral respiratory tract infections, children with RSV were hospitalized longer and received higher respiratory support levels than children with SARS-CoV-2 (**Figure 1C, D, Table 1**). Similarly, children with IV received increased respiratory support than children with SARS-CoV-2 (**Figure 1D, Table 1**). Of the 36 children with SARS-CoV-2, 35 were admitted to the hospital, and 8 required ICU level care. Of those 8 participants, seven were admitted with respiratory symptoms requiring non-invasive respiratory support, while 1 was admitted with severe diabetic ketoacidosis in the setting of fever. None of the SARS-CoV-2-positive children were intubated. However, one patient with a history of obesity (BMI 37.2) met the criteria for pediatric acute respiratory distress syndrome (ARDS) while supported on full-face bilevel positive airway pressure. The age, sex, race, and ethnicity of the children with IV and the uninfected children did not differ from those with SARS-CoV-2 (**Supplemental Figure 1D, F-I**). A larger proportion of children with SARS-CoV-2 identified as Hispanic compared to RSV (**Supplemental Figure 1H**). The sex, race, and ethnicity were not different between uninfected and infected adult groups (**Supplemental Figure 1E-I, Supplemental Table 1**).

**Figure 1.**
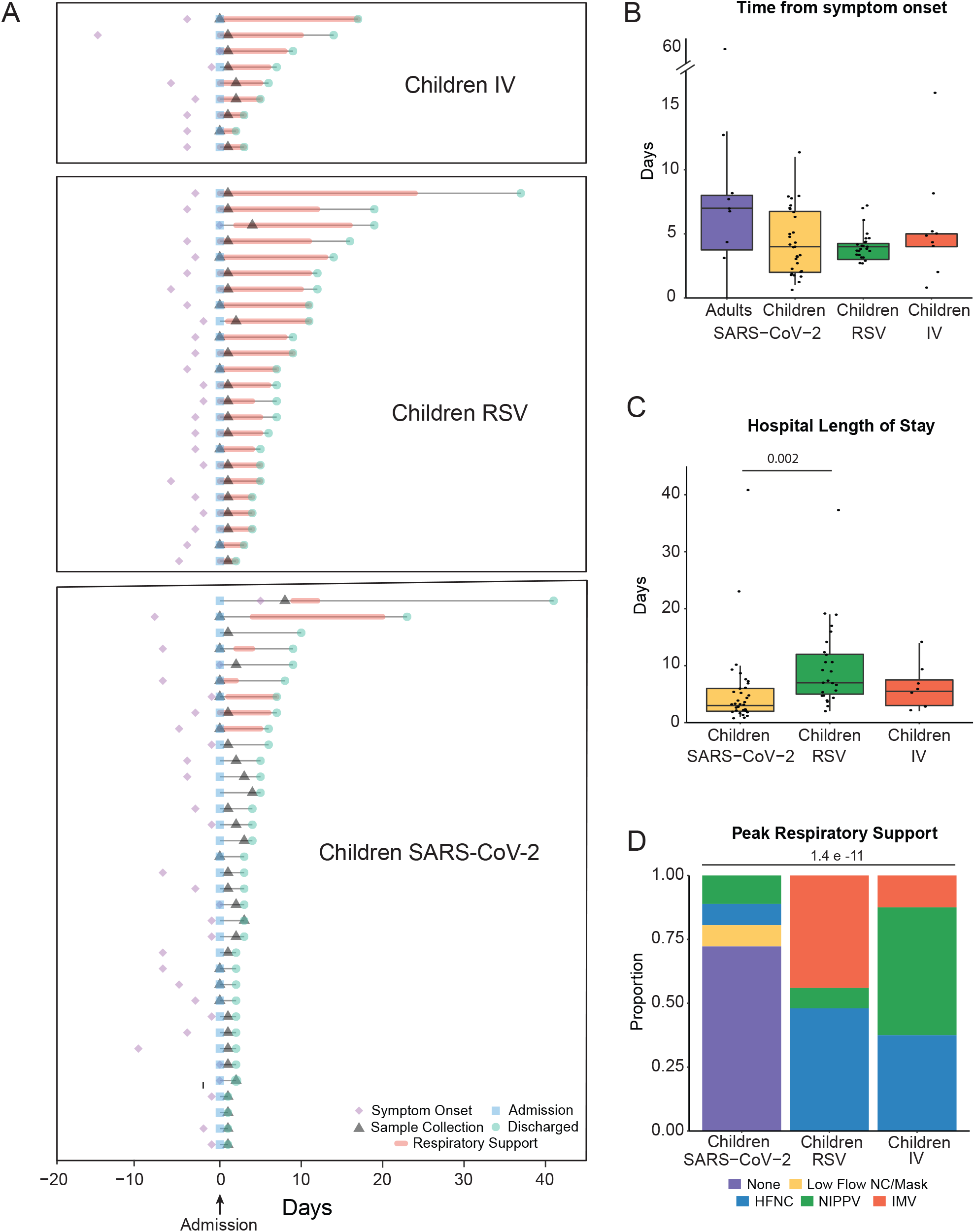
Demographics of the cohort. **A**. Clinical course of pediatric participants. Timing of symptom onset (diamond), admission (square), sample collection (triangle), discharge (circle), hospital stay (thin line), and duration of advanced (HFNC, NIPPV, IMV) respiratory support (bold line) of hospitalized children with SARS-CoV-2 (n=35), RSV (n=25), and IV (n=8). **B**. Self or parent/guardian reported time since symptom onset at the time of sample collection was similar between all participant groups. **C**. Children with RSV infection had a longer hospital length of stay than children with SARS-CoV-2 infection. Pairwise comparisons of medians performed using the Mann-Whitney U test. **D**. Higher proportions of children with IV and RSV infection required HFNC, NIPPV, and IMV at peak illness severity when compared to children with SARS-CoV-2 infection. Proportions compared using Fisher’s Exact Test. All p-values were adjusted using FDR correction. Differences were not significant (p-adjusted>0.05) unless noted. RSV=respiratory syncytial virus, IV=influenza virus, HFNC=high flow nasal cannula, NIPPV=non-invasive positive pressure ventilation, IMV=intermittent mechanical ventilation, FDR=false discovery rate.

The majority of the adult participants with SARS-CoV-2 reported mild illness (**Supplemental Table 2**). There was no difference in the time from symptom onset to nasal sample collection between adults and all pediatric groups (**Figure 1B**). Two adult participants (age 19 and 20 years) were admitted to our pediatric hospital at the time of enrollment. An additional adult participant reported admission to an outside hospital after enrollment in our study. The two patients admitted to our hospital had respiratory illnesses requiring admission to the PICU for non-invasive positive pressure ventilation with supplemental oxygen. Neither met the ARDS criteria. The participant admitted to an outside hospital reported an admission duration of 3 days and did not require intensive care unit level care.

The symptoms reported by participants or the parent/guardian for children with SARS-CoV-2 infection are summarized in **Supplemental Table 1**. Of the 16 adults with SARS-CoV-2 infection (see methods), 7 (44%) were asymptomatic, and the other 9 (56%) reported symptoms. The majority of children with SARS-CoV-2 infection experienced respiratory symptoms (57%). Other symptoms included fever only (14%), gastrointestinal symptoms (9%), non-respiratory viral syndromes (6%), and asymptomatic (14%) (**Table 1**). The asymptomatic children with SARS-CoV-2 were detected on routine screening at admission.

### SARS-CoV-2 viral load and expression of SARS-CoV-2 entry factors in the nasal mucosa does not correlate with age

To characterize our participant sample data and confirm clinical diagnoses, we queried bulk RNA-seq data obtained from curettage of nasal mucosa for the presence of SARS-CoV-2, RSV, or IV reads. Viral reads for SARS-CoV-2 were detected in 28 of 36 samples collected from children diagnosed with SARS-CoV-2 infection by PCR performed in the hospital diagnostic laboratory (**Figure 2A**). Of the 24 children diagnosed with RSV, all had detectable RSV reads in RNA-seq data. Six out of 9 children with IV had detectable Influenza A viral reads in their nasal mucosa. Of the 29 adults accompanying children with SARS-CoV-2 infection, 14 had detectable SARS-CoV-2 reads, and 2 young adults admitted to the pediatric hospital with a positive SARS-CoV-2 PCR test did not have detectable viral reads. Nine samples were obtained from healthy children. RNA-seq did not detect any SARS-CoV-2, RSV, or IV reads in these samples; however, a more in-depth analysis of these samples using the TWIST viral panel as implemented in the OneCodex platform demonstrated the presence of enterovirus and parainfluenza virus in two of the samples. These samples were excluded from subsequent analysis, leaving 7 subjects in the healthy control group (**Supplemental Figure 1C**). In agreement with previous reports (13–16), we found that SARS-CoV-2 viral load was similar between children and adults and did not correlate with *ACE2* expression (**Figure 2B, C**). Several studies have reported that the expression of *ACE2* and *TMPRSS2* – two key entry factors for SARS-CoV-2 – increases with age (11, 12). While some reports suggested that interferons produced in response to viral infection can lead to increased expression of *ACE2*, potentially contributing to the spread of infection (7, 36), other reports have found no correlation or the opposite effect (37–39). We found that the expression of *ACE2* and *TMPRSS2* was not different between healthy children and adults. Moreover, none of the viral infections led to increased expression of *ACE2* and *TMPRSS2* (**Figure 2D**). Similarly, *ACE2* and *TMPRSS2* expression did not correlate with age (**Figure 2E**). Taken together, these data demonstrate that transcriptomic data confirmed clinical diagnoses for SARS-CoV-2, IV, and RSV infections. Moreover, in contrast to previous studies, we did not find a significant correlation between SARS-CoV-2 entry factors and age in our cohort.

**Figure 2.**
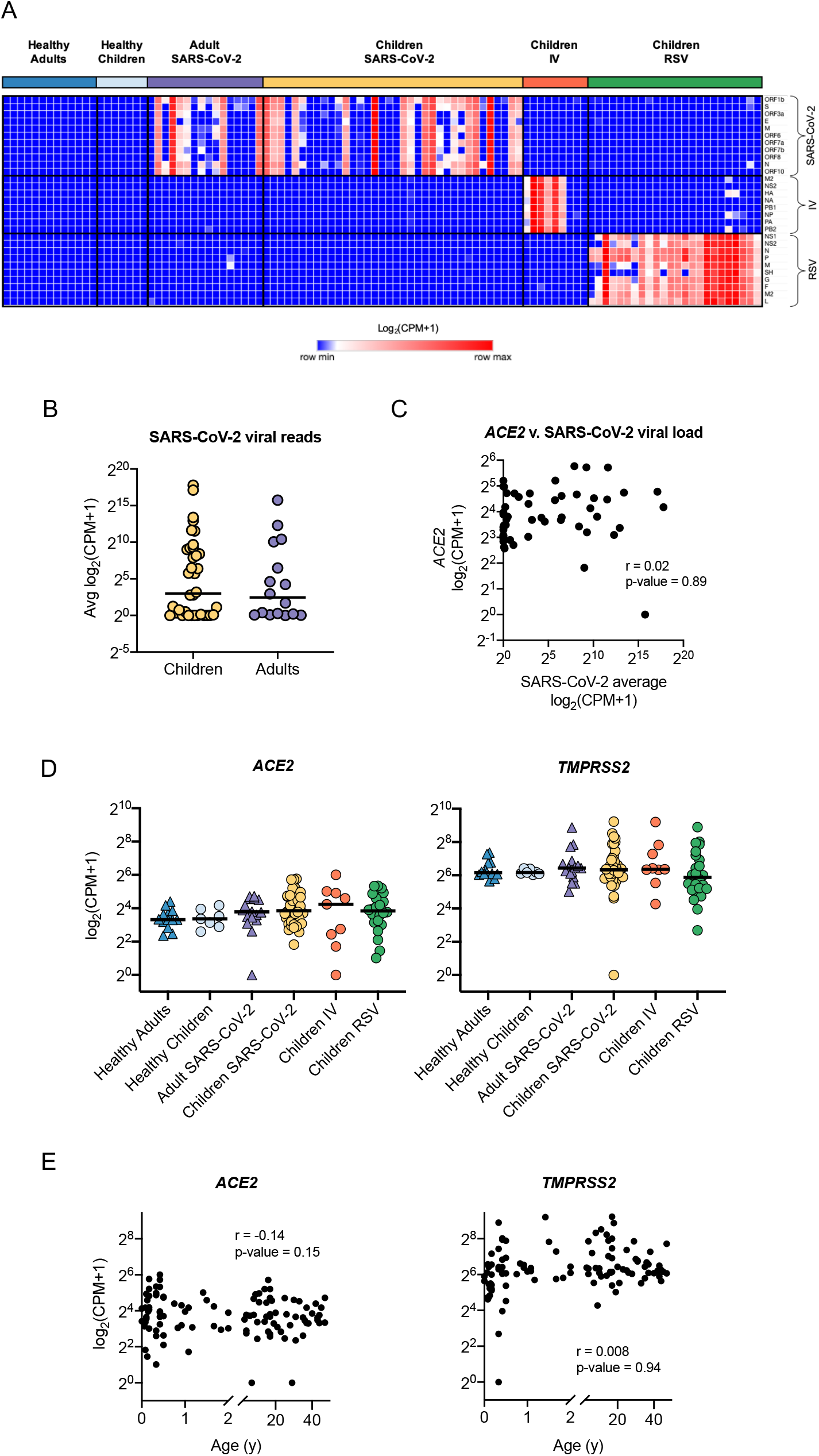
SARS-CoV-2 viral load and expression of SARS-CoV-2 entry factors in the nasal mucosa does not correlate with age. We compared the nasal transcriptome of 36 children with SARS-CoV-2 infection with 7 healthy children, 24 children with RSV infection, 9 children with IV infection, 16 adults with SARS-CoV-2 infection, and 13 healthy adults. **A**. Detection of viral reads for SARS-CoV- 2, influenza A/California/07/2009, and RSV/S2 ts1C in RNA-seq data for all participant samples (n=105). Heatmap of viral genes for SARS-CoV-2, IV, and RSV. Log-transformed pseudocounts (CPM+1) are shown. **B**. Normalized reads were averaged for all 11 SARS-CoV-2 viral genes. Average SARS-CoV-2 reads did not differ between children (yellow, circle) and adults (purple, triangle). Log-transformed pseudocounts (CPM+1) are shown. **C**. Averaged SARS-CoV-2 viral reads did not correlate with *ACE2* expression. Log-transformed pseudocounts (CPM+1) are shown. Pearson correlation coefficient (r) and p-value are shown. **D**. Normalized expression of *ACE2* and *TMPRSS2* was similar across all participant groups; differences not significant, Kruskal-Wallis testing with Benjamini-Hochberg FDR correction, all adjusted p-values >0.05. Log-transformed pseudocounts (CPM+1) are shown. **E**. *ACE2* and *TMPRSS2* expression did not correlate with age. Pearson correlation coefficient (r) and p-value are shown. Log-transformed pseudocounts (CPM+1) are shown.

### Cell-type deconvolution of transcriptomic signatures identifies an influx of immune cells into the nasal mucosa upon viral infection

Bulk RNA-seq data obtained from complex tissue samples composed of multiple cell types can obscure disease-specific signatures as RNA molecules from different cell types are averaged. Accordingly, our initial exploratory analysis did not reveal a distinct structure or pattern within our dataset (**Supplemental Figure 3A, B**). To address the potential heterogeneity in cell-type composition in our samples, we performed *in silico* cell type deconvolution of our bulk RNA-seq data using cell type-specific reference data from the nasal mucosa (35). Hierarchical clustering of predicted cell type abundance distinguished 6 clusters of samples (**Figure 3A, Online Supplemental Table 2**). Four clusters were characterized by an abundance of immune cells (clusters 1–4), while two clusters were enriched with secretory and ciliated epithelial cells (clusters 5 and 6). In line with previous reports (40, 41), deconvolution analysis demonstrated that most nasal mucosa samples from healthy participants were enriched for epithelial cells, with all of the samples from healthy children dominated by a ciliated cell signature (**Figure 3A, B**). In contrast, samples from children with RSV infection were equally divided between the secretory cell cluster and the immune cell clusters, consistent with previous reports of replacement of ciliated cells with secretory cells and the recruitment of neutrophils in RSV infection (42, 43). Increased immune cell abundance was observed in most samples from children infected with IV and SARS-CoV-2 (**Figure 3A, B, Supplemental Figure 3C**). The time from onset of symptoms to the nasal mucosa samples’ collection did not differ between epithelial and immune dominant subsets (**Figure 3C**). In children and adults infected with SARS-CoV-2, viral reads levels were increased in immune-enriched samples compared to epithelial-enriched samples, suggesting increased viral load is associated with immune cell recruitment to the nasal mucosa (**Figure 3D**). To confirm that differences in cell-type composition were not obscuring differences in *ACE2* expression in children and adults, we analyzed *ACE2* expression in samples from the epithelial cell clusters and the immune cell clusters separately. There remained no difference in *ACE2* expression between children and adults **(Figure 3E**). Interestingly, deconvolution analysis of the two samples that were excluded from the healthy children group because of the incidental detection of enterovirus and parainfluenza virus found that these samples had an increased percentage of neutrophils, thus supporting transcriptomic profiling of nasal mucosa as a sensitive tool for assessment of viral infections (**Online Supplemental Table 2**). Taken together, these data demonstrate that samples obtained from the nasal mucosa of healthy and virus-infected participants are characterized by distinct epithelial-or immune-cell abundance profiles. Specifically, samples obtained from participants infected with SARS-CoV-2, IV, or RSV are characterized by an increased abundance of immune cells, whereas healthy participant samples are enriched for epithelial cells.

**Figure 3.**
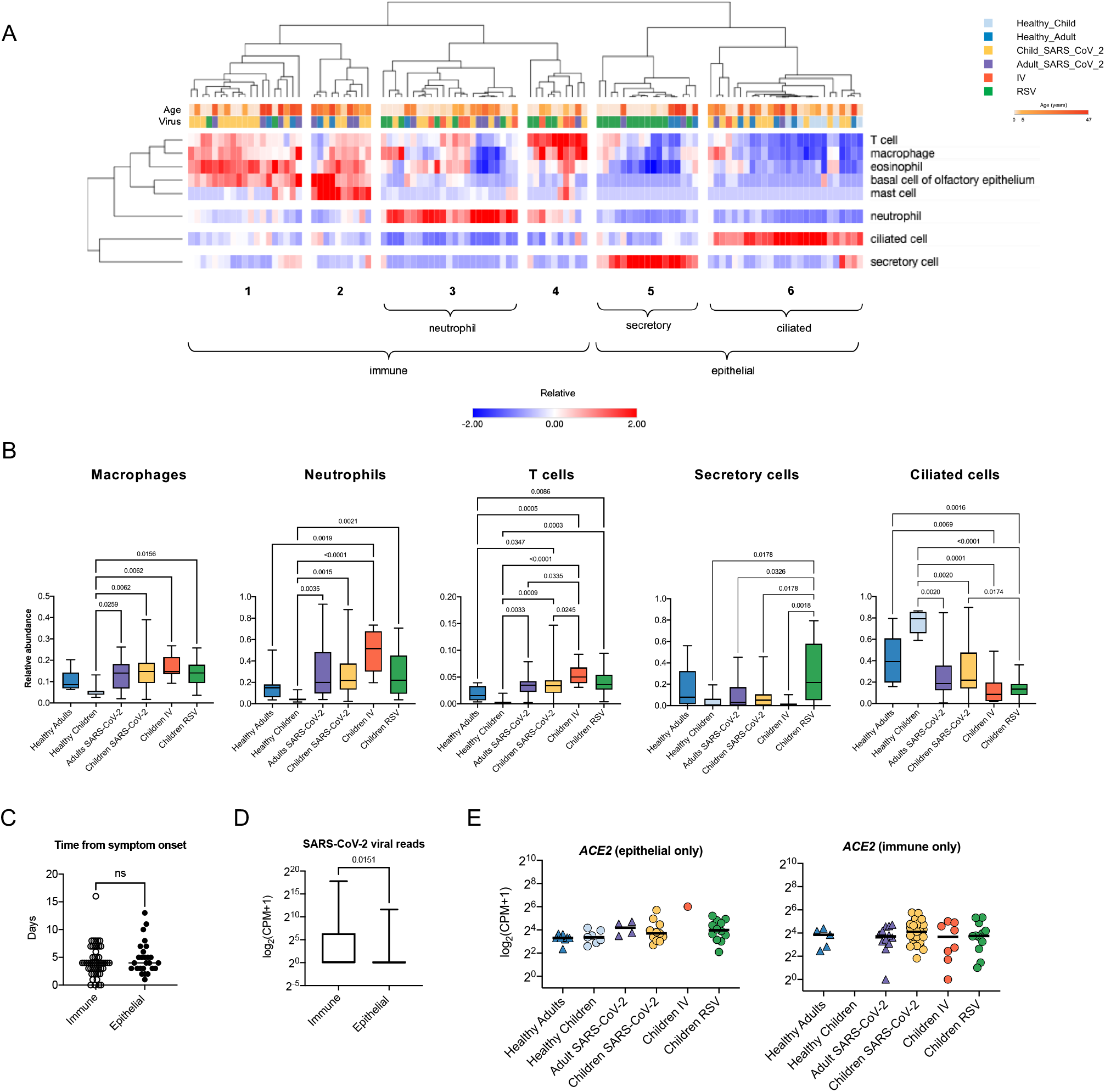
Cell type deconvolution of the transcriptomic signatures in the nasal mucosa demonstrates an influx of immune cells after viral infection. **A**. Heatmap demonstrating the relative abundance of cell types as determined by *in silico* deconvolution of bulk RNA-seq gene expression profiles (see Methods). Hierarchical clustering on samples from 105 participants identified 6 clusters enriched for immune or epithelial cells. Z-score normalized relative abundance values are shown. **B**. Relative abundance of immune and epithelial cell types across all six participant groups. Participants with a viral infection had a significantly higher relative abundance of immune cells than healthy participants. Adjusted p-values calculated using the Kruskal-Wallis test with Benjamini-Hochberg FDR correction are shown. Adjusted p-values <0.05 were considered statistically significant. **C**. Number of days between symptom onset and day of sample collection were similar for immune-enriched and epithelial-enriched samples. n.s.= not significant, by Mann-Whitney U test. **D**. SARS-CoV-2 viral load, defined as average normalized reads for 11 SARS-CoV-2 viral genes, is significantly higher in immune-enriched clusters in comparison to epithelium-enriched clusters. Log-transformed pseudocounts (CPM+1) are shown. The adjusted p-value calculated by the Mann-Whitney U test is shown. **E**. Normalized expression of *ACE2* in epithelial- and immune-enriched samples is similar across all participant groups; differences are not significant, Kruskal-Wallis testing with Benjamini-Hochberg FDR correction, all adjusted p-values >0.05. Log-transformed pseudocounts (CPM+1) are shown.

### Viral infection is associated with increased expression of interferon and innate immune response genes in the nasal mucosa

To investigate whether specific gene expression patterns correlate with clinical parameters, we performed a weighted gene co-expression network analysis (WGCNA). Analysis of all participant samples (n=105, both children and adults) identified 7 modules, 3 of which significantly correlated with one or more clinical parameters **(Figure 4A**). Module 7 positively correlates with age and does not correlate with any other parameter, suggesting it contains age-related genes not associated with a viral infection. Enrichment analysis revealed that this age-related module enriches for genes involved in GO biological processes such as *RNA processing* and includes several small nucleolar RNA genes (snoRNAs) such as *SNORA49* and *SNORD8* (Online Supplemental Table 3), which have been proposed as aging markers (44)(**Figure 4B**). Modules 3 and 4 positively correlated with the number of SARS-CoV-2 viral reads. Module 3, which positively correlated with SARS-CoV-2 and IV reads, enriched for genes involved in *response to virus* and *type I interferon signaling pathway* (**Figure 4B**). Module 4 also correlated with RSV reads and enriched for genes involved in GO biological processes associated with *neutrophil activation, chemotaxis*, and *leukocyte differentiation*.

**Figure 4.**
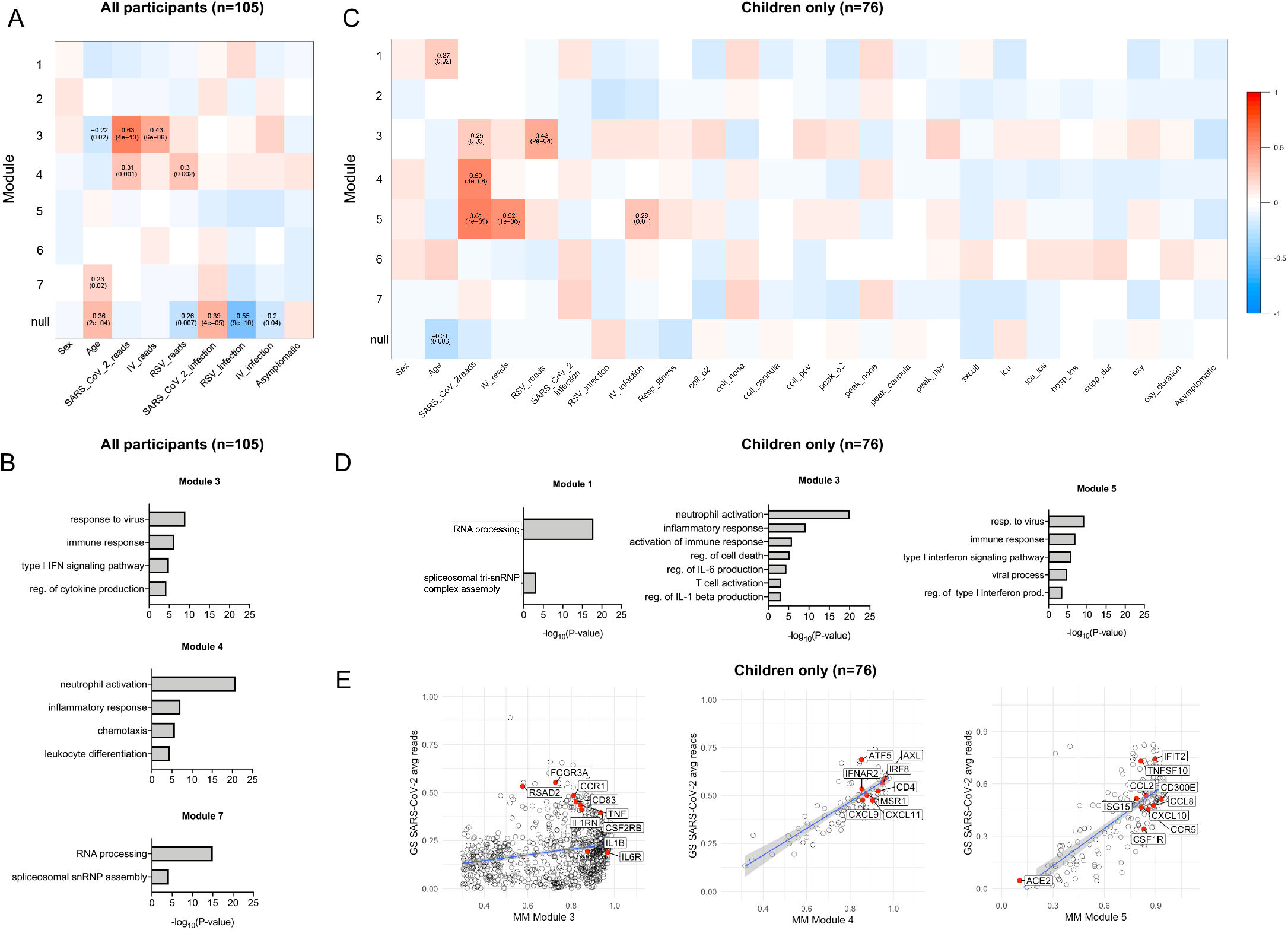
Transcriptional signatures associated with viral infection enrich for inflammatory and interferon response genes in children and adults. **A**. 2170 HVGs for all participant samples (n=105) were analyzed using WGCNA. Module-trait relationships were identified for 7 modules. Modules 1-7 contain 33, 199, 248, 992, 375, 32, and 51 genes, respectively. 240 genes that did not fall into any module were assigned to the null module. **B**. GO biological process enrichment for modules correlating with SARS-CoV-2 viral reads. **C**. 2024 HVGs identified among pediatric participant samples only (n=76) were analyzed using WGCNA. Module-trait relationships were identified for 7 modules. Modules 1-7 contain 43, 385, 823, 92, 200, 204 and 46 genes, respectively. 231 genes that did not fall into any module were assigned to the null module. **D**. GO biological process enrichment for modules correlating with SARS-CoV-2 reads. **E**. Scatterplot of correlation between Gene Significance (GS) for SARS-CoV-2 reads vs. Module Membership (MM) in modules 3,4 and 5. Pearson correlation coefficients (r) and p-values are shown. P-values <0.05 were considered statistically significant. Linear regression line (blue) with a 95% confidence interval (grey) are shown. HVGs - highly variable genes, WGCNA - Weighted Gene Co-Expression Network Analysis. Coll_O2 – On supplemental oxygen at time of collection. Coll_none – No respiratory support at time of sample collection. Coll_cannula – Nasal cannula/simple mask at time of sample collection. Coll_ppv – Positive pressure ventilation (PPV) at time of sample collection. Peak_o2 – Received supplemental oxygen. Peak_none – No respiratory support Peak_cannula – Nasal cannula/simple mask was highest respiratory support during course of illness. Peak_ppv – PPV was highest respiratory support during course of illness. Sx2coll – Time between symptom onset and collection (days). Icu_los – ICU length of stay. Hosp_los – Hospital length of stay. Supp_dur – Duration of support (days). Oxy_duration – Duration of oxygen therapy.

We next focused only on pediatric samples to identify distinct modules correlated with specific viral infections and clinical outcomes. WGCNA identified 7 modules within the pediatric cohort (n=76)(**Figure 4C**). Modules 3, 4, and 5 correlated with SARS-CoV-2 reads (**Figure 4C**). Module 3 also correlated with RSV reads and enriched for genes involved in the GO biological processes *inflammatory response, regulation of IL-6 production*, and *T cell activation* (**Figure 4D**). Module 5 correlated with both SARS-CoV- 2 and IV reads and enriched for genes involved in *response to virus, type I interferon signaling*, and chemokines, including *CXCL10* (**Figure 4E)**. Module 4 is the only module unique to SARS-CoV-2 reads. The 92 genes that make up this module did not enrich for specific GO biological processes but included the antiviral transcription factor *IRF8*, the chemokines *CXCL9* and C*XCL11*, and the T cell marker CD4 (**Figure 4E**). There was no significant correlation between any of the 7 modules and severity of illness or outcomes. Taken together, these data demonstrate that in participants with SARS-CoV-2, RSV, or IV infection, the nasal mucosa is characterized by the expression of genes associated with innate immune and interferon response.

### Local interferon response is independent of age and virus

Since WGCNA identified antiviral interferon signaling as a shared feature of viral infection with SARS-CoV-2, IV, or RSV in our cohort, we next focused on this pathway. We found that gene lists for type I and type II interferon responses largely overlap. Therefore, we compiled a list of IFN-response genes using hallmark pathways for Interferon alpha and gamma responses (45, 46). Comparison of normalized expression across all participant groups revealed an increased expression of interferon response genes in patients with RSV, IV, and SARS-CoV-2 compared to healthy controls that did not correlate with age (**Figure 5A-C**). Specifically, we did not observe differences in the interferon response between children and adults infected with SARS-CoV-2 (**Figure 5B**). While neither age nor type of viral infection correlated with the interferon response, we found a significant correlation with the viral load measured by an average number of viral reads (**Figure 5D**). Consistent with the lack of correlation between viral load, the severity of illness, or the time from symptom onset, the interferon response did not correlate with clinical outcomes such as duration of respiratory support and ICU length of stay (**Supplemental Figure 5A, B**). Interestingly, although we detected an interferon response signature in the majority of virus-infected participants, we did not detect the expression of secreted type I, type II, or type III interferons at the transcriptional level in a majority of patients with RSV or SARS-CoV-2 infection (**Figure 5E**). Secreted interferons were primarily detected in participants with IV infection, consistent with this cohort having the highest average expression of interferon response genes overall (**Figure 5B, E**). This suggests that the nasal mucosa is not a source of secreted interferons in RSV and SARS-CoV-2 infection, at least when these infections are brought to medical attention.

**Figure 5.**
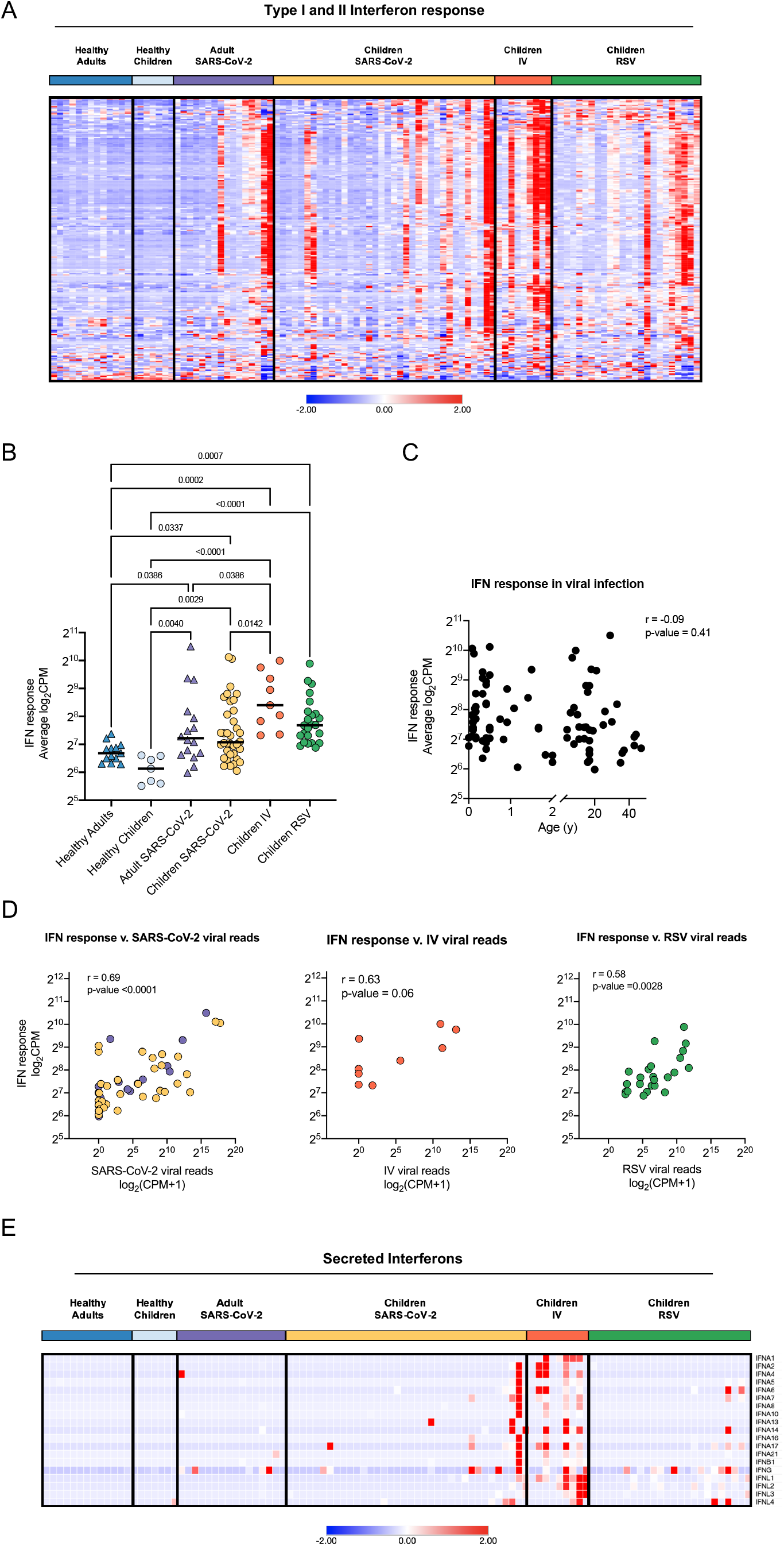
Local interferon response is independent of age and virus. **A**. Heatmap of z-score normalized expression (CPM) of 222 Type I and II IFN response genes. Hierarchical clustering on genes (rows) did not identify distinct clusters. Samples within SARS-CoV-2, IV, and RSV groups are ordered by ascending viral reads for SARS-CoV- 2, IV, or RSV, respectively. **B**. Average normalized expression of IFN response genes (combined Hallmark pathway lists for IFNA and IFNG response) by participant group. Asterisks indicate statistical significance (adjusted p-value <0.05, Kruskal-Wallis test with Benjamini-Hochberg FDR correction). **C**. Scatter plot of correlation between age (years) and average IFN response reads. Log-transformed CPM counts are shown. Pearson correlation coefficient (r) and p-value are shown. **D**. Scatter plot of correlation between average IFN response reads (log_2_CPM) and SARS-CoV-2, RSV, or IV viral loads, respectively. Pearson r-values and p-values are shown. **E**. Heatmap of z-score normalized expression (CPM) for 19 secreted Type I, II, and III interferons.

### Increased expression of innate immune response genes in children with high SARS-CoV-2 viral load

Although IFN response was not different between children and adults infected with SARS-CoV-2 (**Figure 5B**), we wanted to determine if the expression of genes in other pathways was different. Therefore, we next focused only on children and adults with high viral loads (average SARS-CoV-2 viral read > 6 CPM), indicating active infection. We found no difference in the magnitude of the interferon response between children and adults with a high average viral load (**Figure 6A**). The cellular composition was also similar between the groups, allowing us to compare gene expression differences without cellular composition bias (**Figure 6B**). We compared children and adults with high SARS-CoV-2 viral load and identified 576 differentially expressed genes (**Figure 6C**). Genes upregulated in children compared to adults were enriched for innate immune processes, including *cellular response to IL-1* and *inflammatory response* (**Figure 6C, D**). Inflammatory cytokines, their receptors, and chemokines, like *IL18, IL1RL2, CXCL3* and *CCL2*, were among the genes increased in children (**Figure 6C, E**). Participants with low to no SARS-CoV-2 viral load had fewer differentially expressed genes between children and adults and the enrichment analysis did not identify any significant GO biological processes (**Supplemental Figures 6A-B**). Furthermore, we did not observe difference in the IFN response or ACE2 between children and adults with low viral reads (**Supplemental Figures 6B**). There were also no differences in immune or epithelial cellular abundance and no differences in clinical outcomes between low and high viral counts groups (**Supplemental Figures 6C-D**). This data suggests that SARS-CoV-2- infected children with a high viral load have a similar interferon response but an increased innate immune response in the nasal mucosa compared to SARS-CoV-2-infected adults.

**Figure 6.**
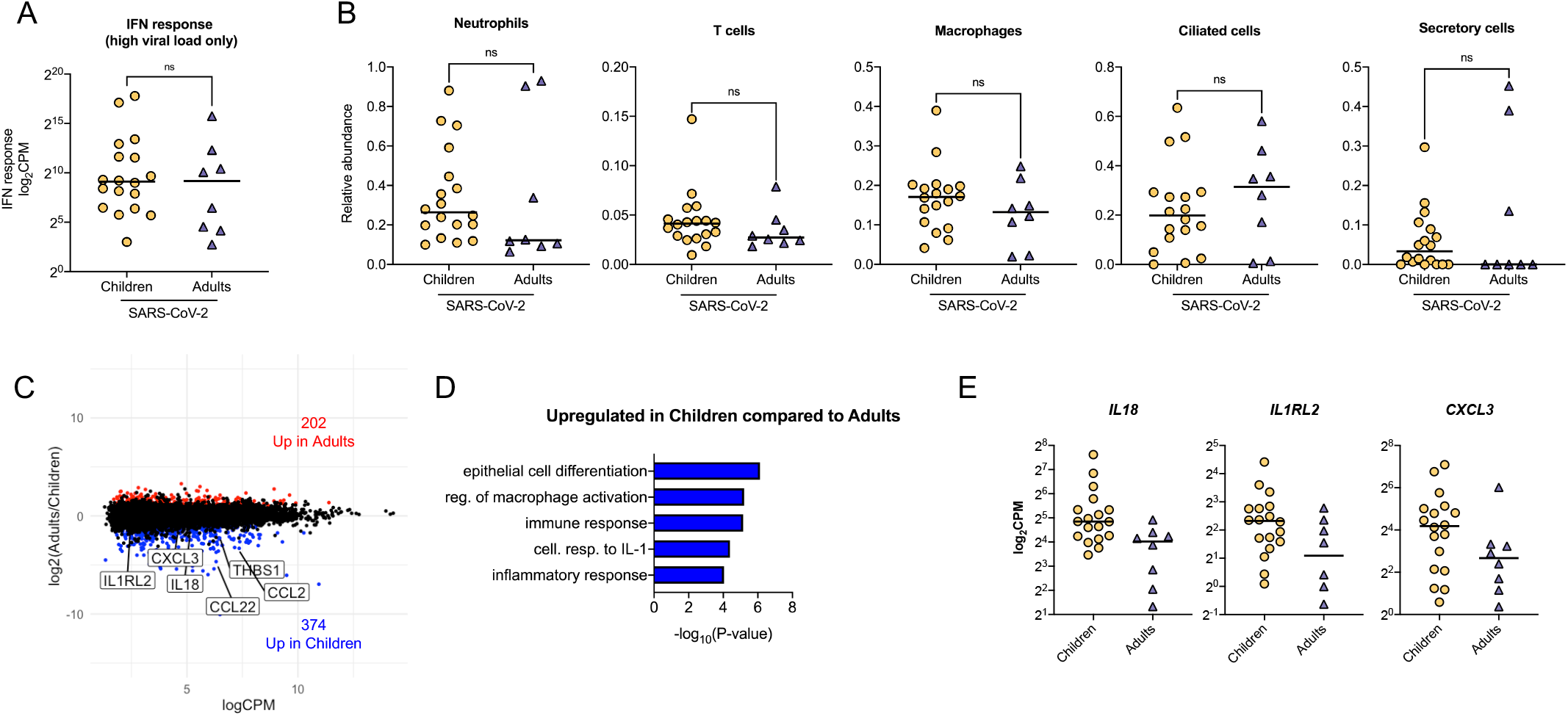
Increased expression of innate immune response genes in children with high SARS-CoV-2 viral load. **A**. The average normalized IFN response gene expression was similar in children and adults with high viral load (>6 CPM average). n.s. = not statistically significant by Kruskal-Wallis test with Benjamini-Hochberg FDR correction. **B**. Relative cellular abundance. n.s. = not significant by Kruskal-Wallis test with Benjamini-Hochberg FDR correction. **C**. MA plot of differentially expressed genes (edgeR p-value <0.05 and log_2_FC > |1.0| between adults and children with high SARS-CoV-2 viral load (> 6 CPM average). Number of genes upregulated (red) and downregulated (blue) in adults compared to children are shown. **D**. Genes upregulated in children enriched for innate immune-related GO biological processes. **E**. Scatter plots of normalized expression for *IL18, IL1RL2*, and *CXCL3* are shown. Line indicates median. Log-transformed CPM counts are shown.

## Discussion

The majority of children infected with SARS-CoV-2 will at most experience mild symptoms similar to common respiratory viral infections. The mechanisms underlying this relative protection from a severe infection in children compared to adults are unclear. Differences in expression of the SARS-CoV-2 entry factors or response to the virus at the primary site of infection between children and adults have been proposed as potential mechanisms (47, 48); however, these hypotheses have not been directly tested to date. Here we present the first study that directly compares host response to SARS-CoV-2 at the primary site of the infection between children and adults. In our cohort of children infected with SARS-CoV-2, the local response in the nasal mucosa was largely the same as non-hospitalized adults infected with SARS-CoV-2 and children with severe RSV and IV infections. This similarity in response to the virus, particularly the interferon response, suggests that age-related differences in SARS-CoV-2 infection outcomes are independent of the primary response to the virus in the nasal mucosa.

Age-related differences in expression of *ACE2* and *TMPRSS2* have been proposed as modulators of clinical severity of SARS-CoV-2 infections. ACE2 is the cell surface receptor bound by the SARS-CoV-2 spike protein, and TMPRSS2 is a host protease that cleaves the spike protein to allow for efficient binding and cell entry (8). Previous studies have demonstrated an association between age and the expression of *ACE2* and *TMPRSS2*, suggesting that low levels of expression of *ACE2* and *TMPRSS2* in pediatric airway epithelial cells could protect from severe infection (11, 12, 36). In our cohort, we did not observe a difference in the expression of *ACE2* or *TMPRSS2* between children and adults. While some reports suggested that *ACE2* expression may increase in response to interferon stimulation (7, 36), we saw no increase in the expression of *ACE2* and *TMPRSS2* expression during active SARS-CoV-2 or other viral infections, supporting previous reports that interferon signaling does not lead to increased *ACE2* expression (37–39).

Our study’s unique strength is the inclusion of the appropriate controls: healthy children and adults and children infected with other respiratory viruses. This allowed us to report our findings on SARS-CoV-2 infection in the context of RSV and IV infections, which are known to induce severe illness requiring hospitalization and respiratory support. We found a high degree of similarity in the local response to SARS-CoV-2, RSV, and IV infections. This agrees with Mick and colleagues’ report, who performed metatranscriptomics on nasopharyngeal swabs and found substantial overlap between SARS-CoV-2 and other acute respiratory infections in adults (49). Thus, our findings suggest that other factors, such as systemic immune responses, may determine the differences in disease severity between children and adults.

Given that the expression of SARS-CoV-2 entry factors and the local response at the primary site of infection does not differ between children and adults, what factors could explain relative protection in children and unpredictable susceptibility in elderly adults infected with SARS-CoV-2? As humans have numerous encounters with betacoronaviruses, including seasonal coronaviruses, they form adaptive immunity (50). Thus, even before the COVID-19 pandemic, a substantial number of individuals in the population had antibodies and T cells that recognize betacoronaviruses and can cross-react with SARS-CoV-2. Such cross-reactive memory T cells have been reported for SARS-CoV (51) and recently for SARS-CoV-2 (52–56). This observation initially led to speculation that these cross-reactive antibodies and T cells could play a protective role in children and young adults and that vanishing adaptive immunity would make elderly individuals more susceptible to SARS-CoV-2. However, in another study, a larger number of SARS-CoV-2 cross-reactive memory T cells was observed independently in the elderly and patients with severe compared with mild COVID-19 (56). Importantly, these cross-reactive T cells exhibited lower avidity and reduced antiviral responses when stimulated with SARS-CoV-2 peptides compared with T cells from patients who recovered from COVID-19. Thus, instead of being protective, these cross-reactive low avidity T cells, which are less efficient at virus elimination, could contribute to prolonged production of inflammatory signaling. This hypothesis is attractive as it explains the differences in the epidemiology of SARS-CoV-2, which disproportionately affects elderly individuals. Children and young adults, who likely have fewer lifetime coronavirus exposures, often have only mild symptoms, despite having viral loads in the upper airways comparable to adults (3, 14). While multiple studies have investigated adaptive immune response in adults infected with COVID-19, similar systematic studies in the pediatric population are lacking.

Our study has several limitations. First, our institution utilized two different PCR platforms for the detection of SARS-CoV-2 during the study period. Due to differences in methodology, viral loads cannot be directly compared between the platforms. Moreover, PCR testing was not available for the adults accompanying children infected with SARS-CoV-2 during the study period. To overcome this limitation, we used dual RNA-seq, which allows for simultaneous host transcriptomic analysis and pathogen characterization (21). By creating custom hybrid genomes, we were able to identify and quantify viral transcripts in our samples as a surrogate for viral load, thus, enabling comparison across all groups in our study. Second, while our study includes children infected with other respiratory viruses – RSV and IV, which serve as proper controls to SARS-CoV-2-infected children, it primarily includes hospitalized children with SARS-CoV-2, representing a minority of pediatric SARS-CoV-2 infections. However, 14% of our pediatric cohort were asymptomatic with incidental SARS-CoV-2 diagnoses. The local host response in these asymptomatic children’s nasal mucosa could not be distinguished from more severely affected pediatric participants. Similarly, our study only included two hospitalized young adults with moderate SARS-CoV-2 infection and did not include any severely ill hospitalized adults requiring intubation and mechanical ventilation. Therefore, hospitalized adults with severe SARS-CoV-2 may have a different local response at the primary site of infection. However, the high degree of similarity in the local response between different respiratory viruses in our cohort suggests that other factors, such as the systemic immune response, determine disease severity in SARS-CoV-2 infection. Supporting this interpretation, recent studies using single-cell RNA-seq profiling of nasopharyngeal swabs demonstrated activation of an interferon-response signature in epithelial cells and influx of immune cells in hospitalized adults with severe COVID-19 (57, 58). Third, our transcriptomic analysis was performed on a biopsy from a complex tissue containing multiple epithelial and immune cell types (40). Thus, our results are affected by the changes in the abundance of the specific cell types, either due to age- or disease-related differences in tissue composition or operator sampling bias. We applied *in silico* tissue deconvolution (34) to account for differences in tissue composition, which allowed us to perform a more accurate comparison between samples with similar composition. This comparison demonstrated that among subjects with high SARS-CoV- 2 viral load, children exhibit increased activation of the innate immune system on a per-cell level. Ultimately, the use of techniques that enable analysis at the single-cell level, such as single-cell RNA-seq, will allow a more precise assessment of cell-type-specific changes associated with SARS-CoV-2 or other viral infections. Recently, single-cell RNA-seq has been applied to understand virus-host interactions in children infected with IV (59), and similar efforts are underway in SARS-CoV-2 (60).

In summary, we present evidence that the response to SARS-CoV-2 infection at the primary site of infection is similar between children and adults and comparable in its magnitude to other common respiratory viruses. Our findings suggest that factors outside of the primary infection site are likely drivers of the differences in severity of SARS-CoV- 2 infection between children and adults.

## Data Availability

Processed RNA-seq counts are available as Online Supplemental Table 1. Raw data is
in the process of being deposited to dbGaP/SRA. Code is available via
https://github.com/NUPulmonary/2021_Koch.

## Acknowledgments and funding

This research was supported in part through the computational resources and staff contributions provided by the Genomics Compute Cluster, which is jointly supported by the Feinberg School of Medicine, the Center for Genetic Medicine, and Feinberg’s Department of Biochemistry and Molecular Genetics, the Office of the Provost, the Office for Research, and Northwestern Information Technology. The Genomics Compute Cluster is part of Quest, Northwestern University’s high-performance computing facility, to advance research in genomics.

The authors thank all the participants for enrolling in this study, The authors Rogan Grant for his help with the RNA-seq pipeline and Benjamin Singer and G.R.S. Budinger for their thoughtful discussions.

Andrew D. Prigge was supported by the Gorter Family Foundation. Alexander V. Misharin was supported by N.I.H. grants U19AI135964, P01AG049665, R01HL153312, and NUCATS COVID-19 Rapid Response Grant. Karen M. Ridge was supported by N.I.H. grants P01AG049665 and P01GM096971. Bria M. Coates was supported by funds provided by the Manne Research Institute COVID-19 Springboard Exploratory Research Award and N.I.H. grant K08HL143127. Taylor Heald-Sargent and L. Nelson Sanchez-Pinto were supported by funds provided by the Manne Research Institute COVID-19 Springboard Exploratory Research Award.

The authors do not declare a conflict of interest.

## Figure and Table Legends

**Supplemental Figure 1.**
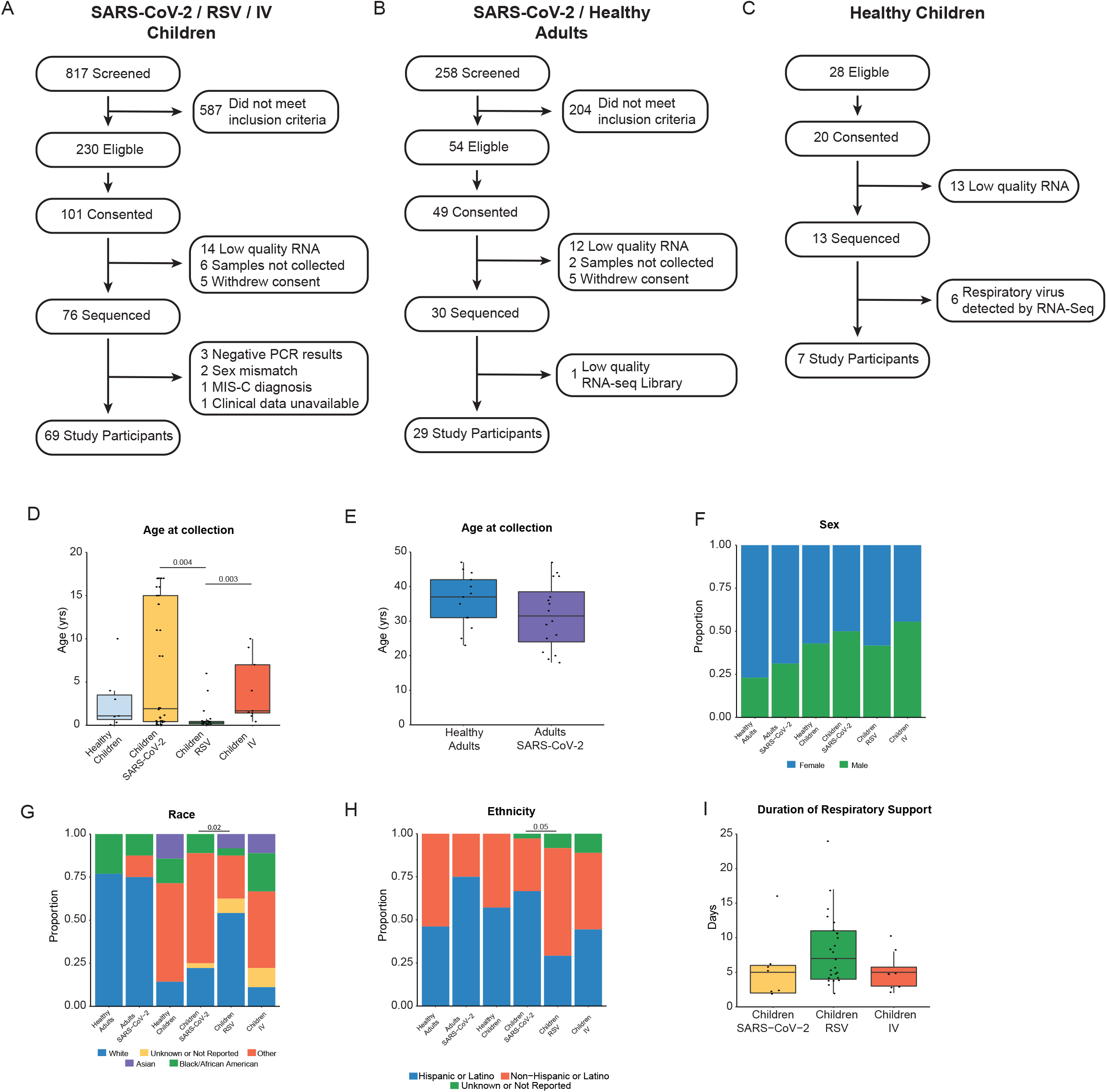
Flow chart of study subject recruitment and sample analysis. **A**. Enrollment of children with RSV, IV, and SARS-CoV-2 infection from December 2019 to November 2020. **B**. Enrollment of adults accompanying children with SARS-CoV-2 infection and young adults with SARS-CoV-2 infection admitted to our hospital from March 2020 to November 2020. **C**. Enrollment of healthy children in a previous study in May 2018. **D**. Children with RSV infection were younger than those with SARS-CoV-2 and IV infections. **E**. Distribution of ages between adults infected with SARS-CoV-2 and healthy adults was similar. **F**. Proportion of females (blue) and males (green) were similar between all groups. **G**. Proportions of self or parent/guardian reported race differed between children with SARS-CoV-2 and RSV infections. **H**. A larger proportion of children with SARS-CoV-2 infection identified as Hispanic or Latino when compared to children with RSV infection. **I**. Among children requiring advanced respiratory support, there was no difference in the duration of respiratory support. Pairwise comparisons of medians calculated using the Mann-Whitney U test. Proportions compared using Fisher’s Exact Test. All p-values were adjusted using FDR correction. Differences were not significant (p-adjusted>0.05) unless noted. RSV=respiratory syncytial virus, IV=influenza virus, HFNC=high flow nasal cannula, NIPPV=non-invasive positive pressure ventilation, IMV=intermittent mechanical ventilation, FDR=false discovery rate.

**Supplemental Figure 3.**
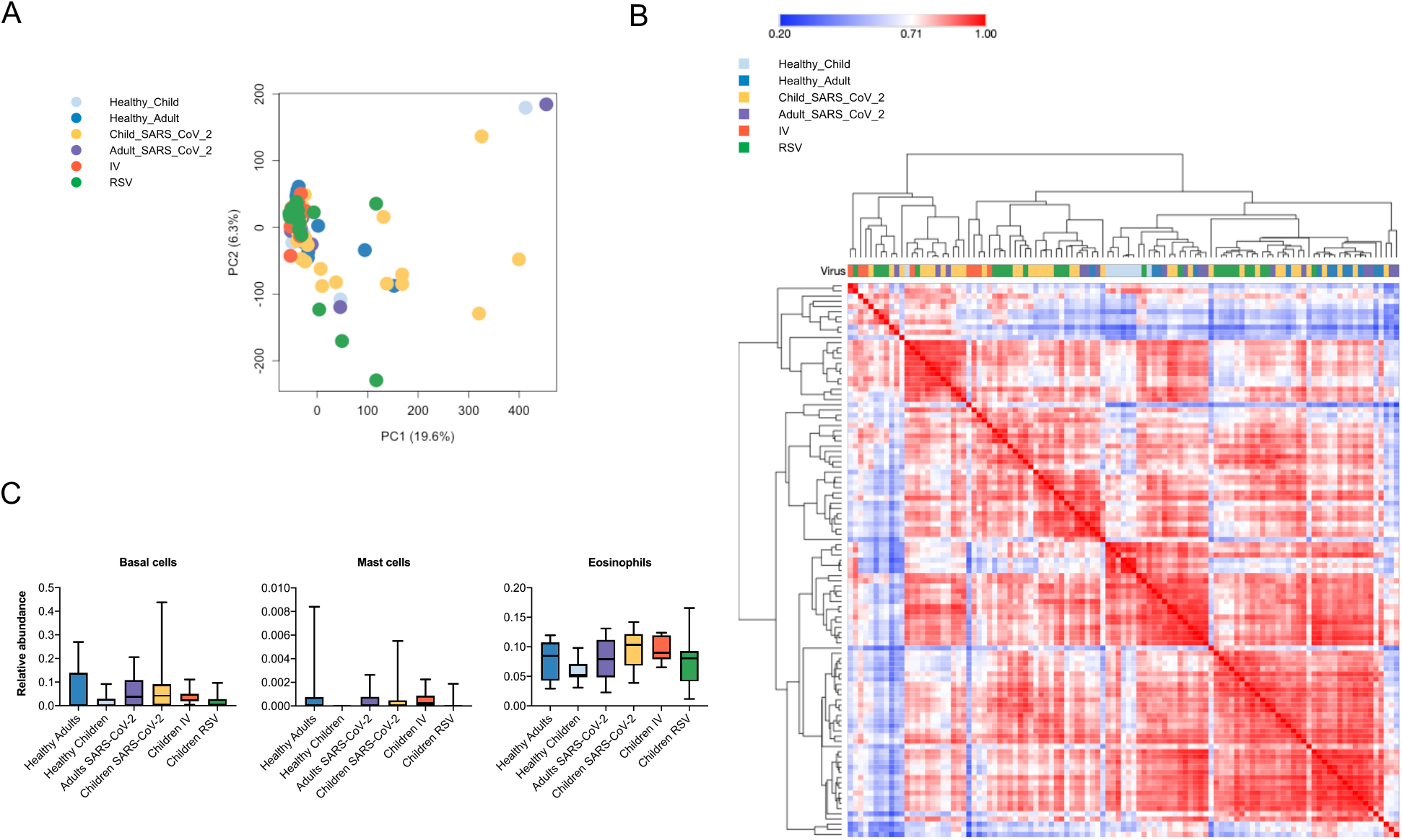
**A**. Principal component analysis (PCA) of normalized counts data (CPM) from all participant samples (n=105) demonstrates a lack of structure related to age or diagnosis. **B**. Heatmap of Pearson correlation of mean gene expression across all samples, hierarchical clustering performed on rows and columns. Participant samples do not cluster by age or diagnosis, except for the healthy children group. **C**. Relative abundance of immune and epithelial cell types across all six participant groups. Adjusted p-values were calculated by Kruskal-Wallis with Benjamini-Hochberg FDR correction.

**Supplemental Figure 5.**
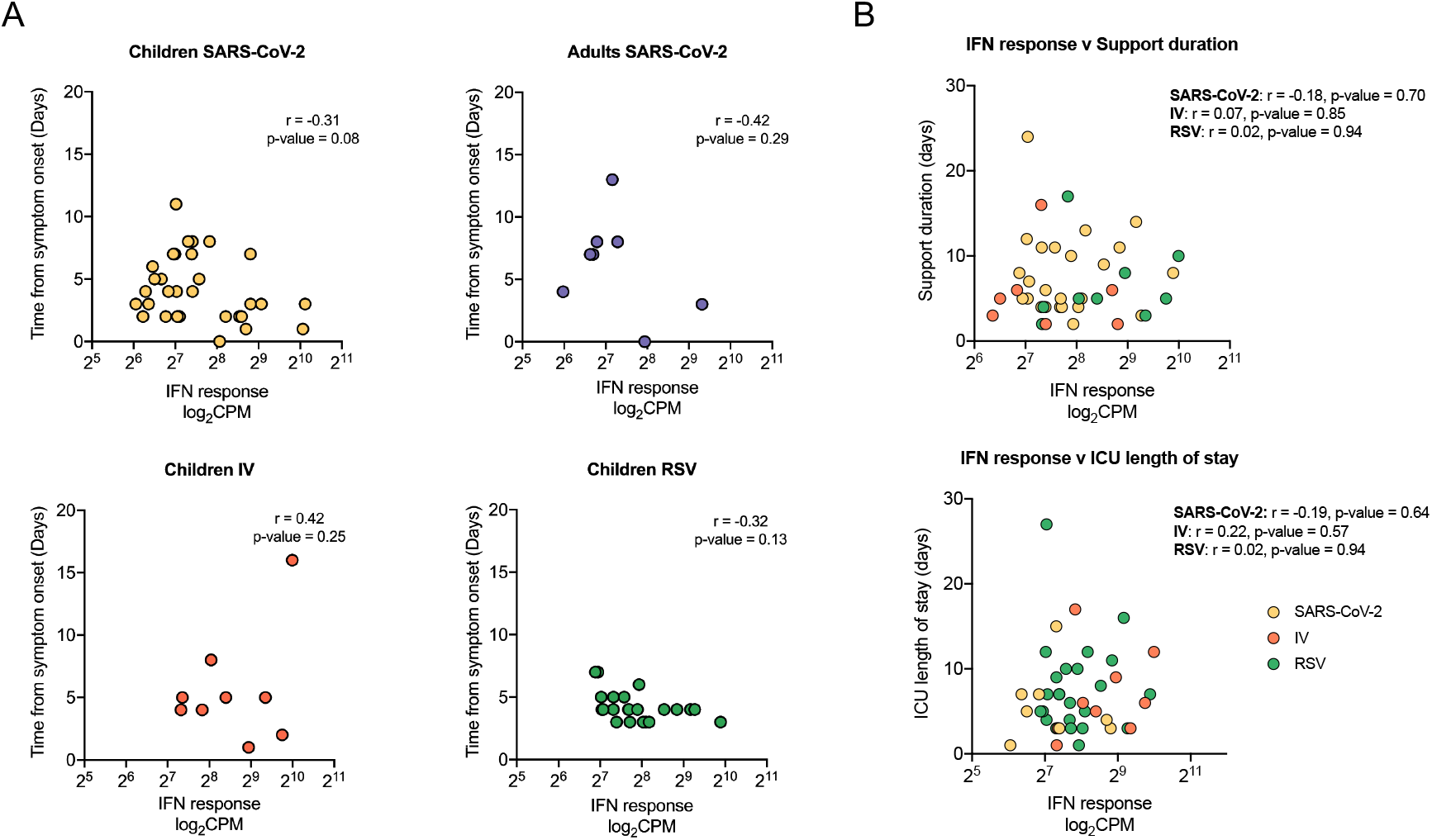
**A**. Time from symptom onset to day of collection in children and adults with SARS-CoV-2, RSV, or IV infection. Pearson correlation coefficient (r) and p-value are shown. **B**. Scatter plot of correlation between support duration (days) or ICU length of stay compared to average IFN response. Pearson correlation coefficient (r) and p-value are shown.

**Supplemental Figure 6.**
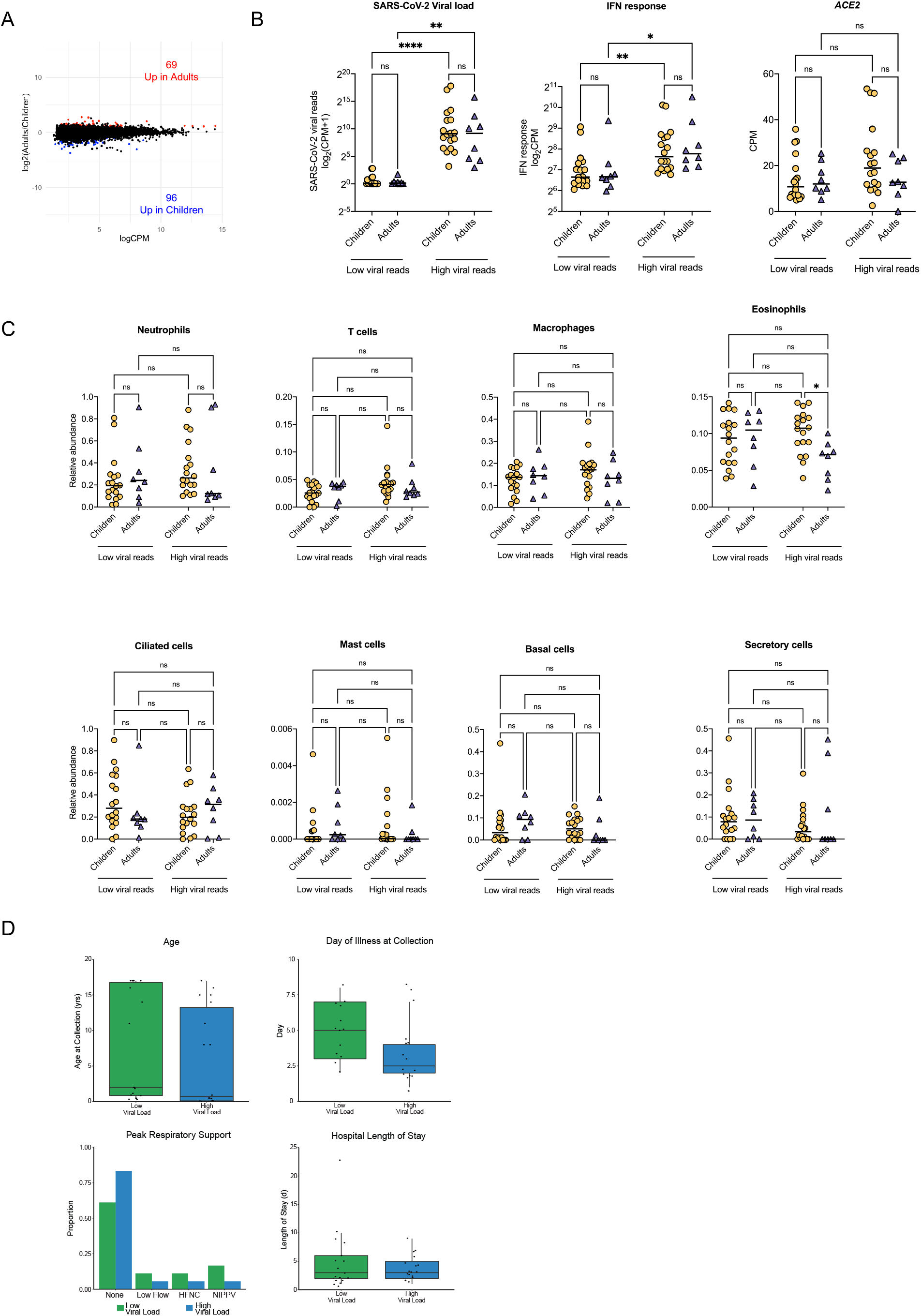
**A**. MA plot of differentially expressed genes (edgeR p-value <0.05 and log_2_FC > |1.0| between adults and children with low to no SARS-CoV-2 viral load (< 6 CPM average). Number of genes upregulated (red) and downregulated (blue) in adults compared to children are shown. **B**. SARS-CoV-2 viral load, defined as average normalized reads for 11 SARS-CoV-2 viral genes, is similar between children and adults in the low and high viral reads groups. Log-transformed pseudocounts (CPM+1) are shown. The average normalized IFN response gene expression was similar between children and adults with low (< 6 CPM average) or high viral load (> 6 CPM average). *ACE2* expression did not differ between children and adults with low (< 6 CPM average) or high viral load (> 6 CPM average). n.s. = not statistically significant by Kruskal-Wallis test with Benjamini-Hochberg FDR correction. **C**. Relative cellular abundance. n.s. = not significant by Kruskal-Wallis test with Benjamini-Hochberg FDR correction. **D**. No difference in clinical characteristics of pediatric participants with low (< 6 CPM average) or high viral load (> 6 CPM average).

**Supplemental Table 1:** Description of Adult Cohort. Adjusted p-value <0.05 were considered statistically significant, calculated by Kruskal-Wallis test with Benjamini-Hochberg FDR correction.

|  | Healthy Adults | SARS-CoV-2 Adults | P-Adj |
| --- | --- | --- | --- |
| **n** | 13 | 16 |  |
| **Median Age (yrs)** | 37 [31-42] | 31.5 [24.0-38.5] | 0.48 |
| **[IQR] Sex** |  |  |  |
| Male (%) | 3 (23.1) | 5 (31.3) | 0.89 |
| Female (%) | 10 (76.9) | 11 (68.8) |  |
| **Race** |  |  |  |
| White (%) | 10 (76.9) | 12 (75.0) | 0.74 |
| African American (%) | 3 (23.1) | 2 (12.5) |  |
| Asian (%) | 0 (0.0) | 2 (12.5) |  |
| Other (%) | 0 (0.0) | 0 (0.0) |  |
| Unspecified (%) | 0 (0.0) | 0 (0.0) |  |
| **Ethnicity** |  |  |  |
| Non-Hispanic (%) | 7 (53.8) | 4 (25.0) | 0.38 |
| Hispanic (%) | 6 (46.2) | 12 (75.0) |  |
| Unspecified (%) | 0 (0.0) | 0 (0.0) |  |
| **Medical History** |  |  |  |
| Healthy (%) Respiratory | 10 (62.5) | 10 (76.9) | 0.74 |
| (%) Cardiovascular (%) | 4 (25.0) | 0 (0.0) | 0.32 |
| Oncologic (%) | 1 (6.25) | 1 (7.7) | 1 |
| Endocrine (%) | 1 (6.25) | 0 (0.0) | 1 |
|  | 2 (12.5) | 3 (23.1) | 0.88 |

**Supplemental Table 2:**
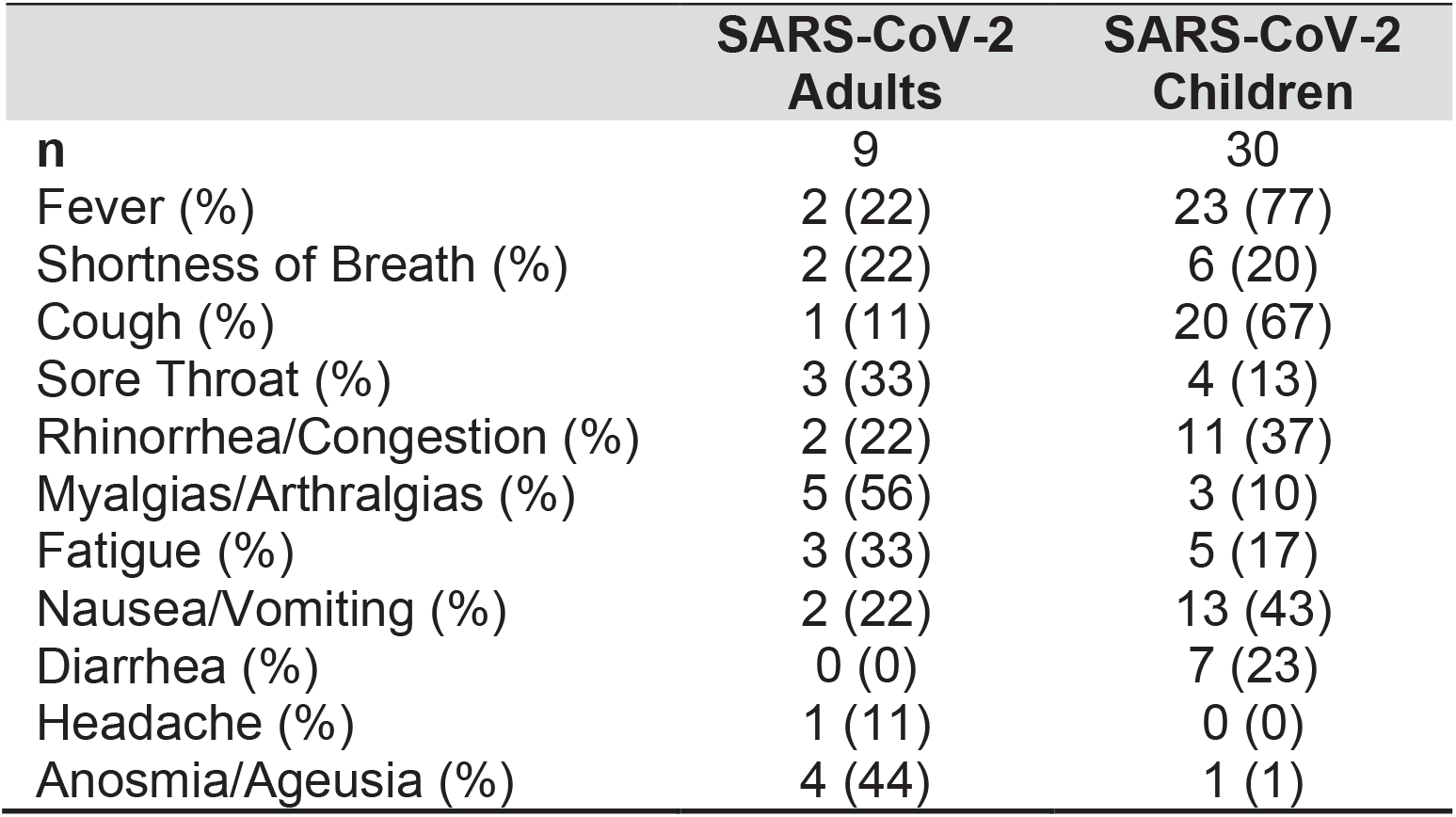
Self or parent/guardian reported SARS-CoV-2 symptoms.

|  | SARS-CoV-2 Adults | SARS-CoV-2 Children |
| --- | --- | --- |
| **n** | 9 | 30 |
| Fever (%) | 2 (22) | 23 (77) |
| Shortness of Breath (%) | 2 (22) | 6 (20) |
| Cough (%) | 1 (11) | 20 (67) |
| Sore Throat (%) | 3 (33) | 4 (13) |
| Rhinorrhea/Congestion (%) | 2 (22) | 11 (37) |
| Myalgias/Arthralgias (%) | 5 (56) | 3 (10) |
| Fatigue (%) | 3 (33) | 5 (17) |
| Nausea/Vomiting (%) | 2 (22) | 13 (43) |
| Diarrhea (%) | 0 (0) | 7 (23) |
| Headache (%) | 1 (11) | 0 (0) |
| Anosmia/Ageusia (%) | 4 (44) | 1 (1) |

